# SARS-CoV-2 sequencing reveals rapid transmission from college student clusters resulting in morbidity and deaths in vulnerable populations

**DOI:** 10.1101/2020.10.12.20210294

**Authors:** Craig S. Richmond, Arick P. Sabin, Dean A. Jobe, Steven D. Lovrich, Paraic A. Kenny

## Abstract

College reopening decisions during the SARS-CoV-2 pandemic represent a trade-off between competing risks to students, faculty and staff, and college finances. Additionally, risks taken in reopening colleges can impose significant burdens on individuals living in surrounding communities. Many colleges that reopened for in-person instruction have reported frequent SARS-CoV-2 outbreaks. La Crosse County, Wisconsin experienced a substantial SARS-CoV-2 outbreak (2,002 cases in September 2020) that coincided with the return to in-person instruction at three local academic institutions. Genomic sequencing of SARS-CoV-2 cases in La Crosse during that period found rapid expansion of two viral substrains. Although the majority of cases were among college-age individuals, from a total of 111 genomes sequenced we identified rapid transmission of the virus into more vulnerable populations. Eight sampled genomes represented two independent transmission events into two skilled nursing facilities, resulting in two fatalities. Our study highlights the very significant risks imposed by college administrator reopening decisions, not just on college-associated populations, but on vulnerable individuals in surrounding communities.

## INTRODUCTION

The SARS-CoV-2 betacoronavirus is a novel respiratory pathogen which emerged in Wuhan, China in late 2019 (1). Numerous teams around the world have produced a rapidly increasing number of publicly available sequences (116,650 in GISAID on 9/28/20) with obvious sequence diversification as viral replication and spread has continued. These sequence variants, inherited in each cycle of viral replication, allow the generation of phylogenetic trees through which the evolution of the virus can be traced. Individual viruses acquire mutations during replication at an approximate rate of 1-2 sites per month (2). This mutation rate provides good resolution for both global and local tracking of viral substrains. Because viral replication occurs in individuals, tracking these inherited variants acts as a proxy for tracking the spread of the virus through populations of individuals.

Starting in late March 2020, we have monitored the introduction and spread of SARS-CoV-2 to the service area of the Gundersen Health System, an integrated healthcare system headquartered in La Crosse, WI and providing care in 21 counties in southwestern Wisconsin, northeastern Iowa and southeastern Minnesota. We have performed whole viral genome sequencing on 514 positive cases from this region, spanning three states which have adopted mitigation approaches of differing intensity.

In September 2020, La Crosse County (Wisconsin) experienced a rapid increase in cases, strongly driven by infections among the 10-19 and 20-29 age groups. The period coincided with the return of college students to three institutions of higher education in the city of La Crosse: University of Wisconsin La Crosse, Western Technical College and Viterbo University, with total enrollments of 10,558, 4,004 and 2,592, respectively (3). Our regional SARS-CoV-2 surveillance and sequencing program captured many of these genomes, allowing assessment of the substrain contributions to the outbreak and analysis of the speed with which vulnerable non-student populations were affected.

## MATERIALS AND METHODS

### Specimens

Specimens were nasopharyngeal swabs taken for diagnostic purposes and evaluated for SARS-CoV-2 positivity at the Gundersen Medical Foundation’s Molecular Diagnostics Laboratory using the CDC 2019-nCoV Real-Time RT-PCR Diagnostic assay. This study includes positive cases diagnosed between 3/18/2020 and 9/23/2020. Ethical approval was obtained from the Gundersen Health System Institutional Review Board (#2-20-03-008; PI: Kenny) to perform additional next-generation sequencing on remnant specimens after completion of diagnostic testing. Samples for this study include only those testing positive in Gundersen Healthcare System’s diagnostic laboratories and, as such, do not include other cases from the region which may have been tested in other healthcare systems and/or public health laboratories.

### RNA isolation and cDNA synthesis

Nasopharyngeal swabs were immersed in liquid medium. RNA was isolated using QIAmp Viral RNA Mini kit (Qiagen, Germantown, MD). cDNA was synthesized from 5 μl of purified RNA using ProtoScript II First Strand cDNA Synthesis Kit (New England Biolabs, Ipswich, MA) with random hexamers.

### Next Generation sequencing

We used the Ion AmpliSeq SARS-CoV-2 Panel (Thermo-Fisher, Waltham, MA) to sequence 237 viral specific targets encompassing the complete viral genome from cDNA derived from SARS-COV-2-positive clinical specimens. Barcoded multiplexed libraries were prepared in batches of eight on the Ion Chef (Thermo-Fisher) and sequenced on the Ion Torrent S5 (Thermo-Fisher).

### Bioinformatics

Demultiplexed Tmap-aligned bam files were produced by the Ion Torrent S5. To overcome difficulties caused by read soft-clipping by this non-splice aware aligner, individual sequence reads were extracted using samtools (4) and re-aligned to the SARS-CoV-2 genome using a splice-aware aligner, hisat2 (5). These bam files were subjected to QC review in IGV. Samples passing QC had a consensus sequence computed according to the method of Cavener (6). Consensus sequences were reported to GISAID (7). For phylogenetic inference (i.e. to determine the hierarchy of case relationships) samples were integrated with associated metadata and aligned on a local implementation of NextStrain (8) using augur and displayed via a web browser using auspice. Choropleth maps were generated using folium.

## RESULTS

The cumulative history of SARS-CoV-2 cases, stratified by age, for La Crosse county is shown in Figure 1. Notable events along this timeline include the reopening of bars in Wisconsin (5/14/2020) and the onset of warm summer weather (first day above 80’F was on Memorial Day Weekend on 5/24/2020) which preceded a rapid rise in cases in early-mid June. Case growth continued at a slower rate for much of the summer before abruptly increasing in the second week of September, coinciding with the return of students to three colleges located in the city of La Crosse. Analysis of cases by census tract showed that the most rapid case growth was occurring the immediate vicinity of the three colleges, an area of high density student housing (Figure 2).

**Figure 1.**
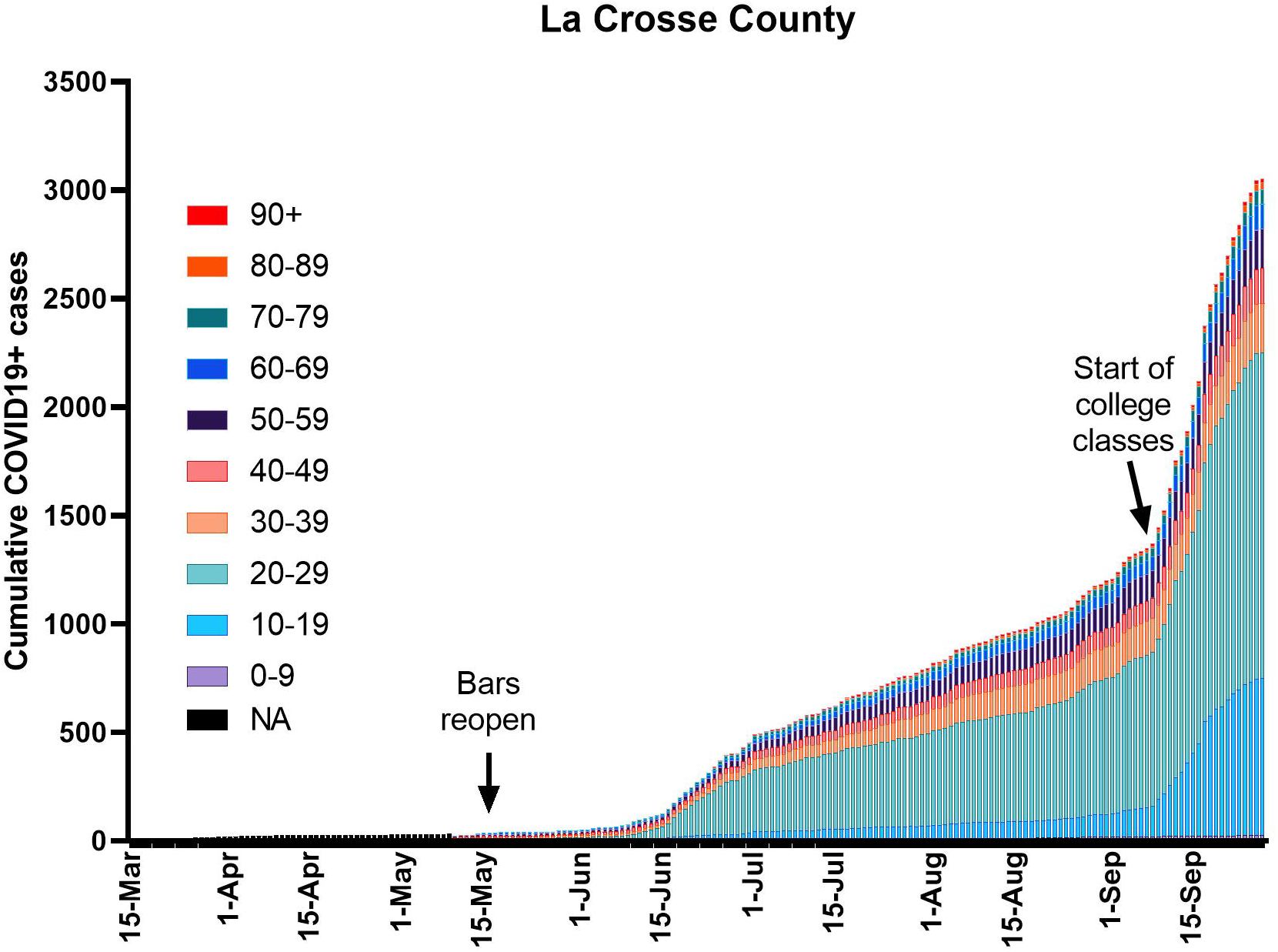
Cumulative SARS-CoV-2 case incidence in La Crosse County, WI, stratified by age group (March – September 2020).

**Figure 2.**
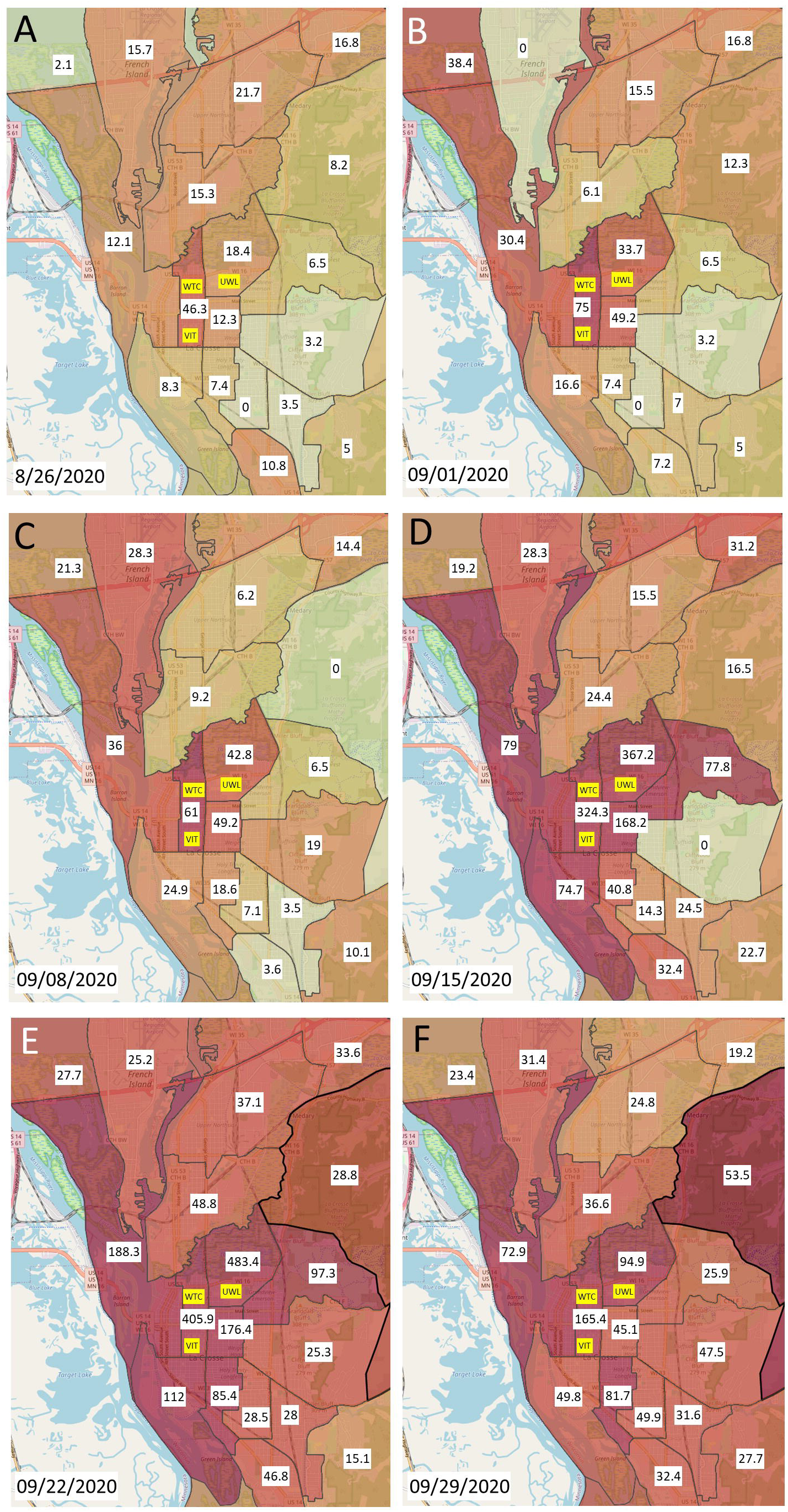
Choropleth heatmaps showing a weekly assessment of the 7-day rolling average of daily case numbers per 100,000 population in census tracts in the city of La Crosse. Census tract populations were taken from 2010 census data. The locations of University of Wisconsin – La Crosse (UWL), Western Technical College (WTC) and Viterbo University (VIT) are indicated.

Between 3/18/20 and 9/23/20 we obtained 514 high quality genomes (Figure 3A) from surveillance in a 21-county region of western Wisconsin, southeastern Minnesota and northeastern Iowa. Clades are colored for ease of visualization and include several epidemiologically relevant outbreaks. Significant examples include (i) an outbreak originating in a meatpacking plant in Postville, IA (9) and (ii) a large outbreak associated with a cluster of bars in downtown La Crosse in June/July. Figure 3B highlights newly sequenced genomes in our region in September 2020. The overwhelming majority of case growth in La Crosse county was explained by two clusters, designated “College A” and “College B”. The small light-blue cluster at the 11 o’clock position represents a substrain detected in the city of Holmen, which did not involve college students, while the other highlighted clades represented case growth in our region outside of La Crosse county. Here we focus on the analysis of the College A and College B clusters.

**Figure 3.**
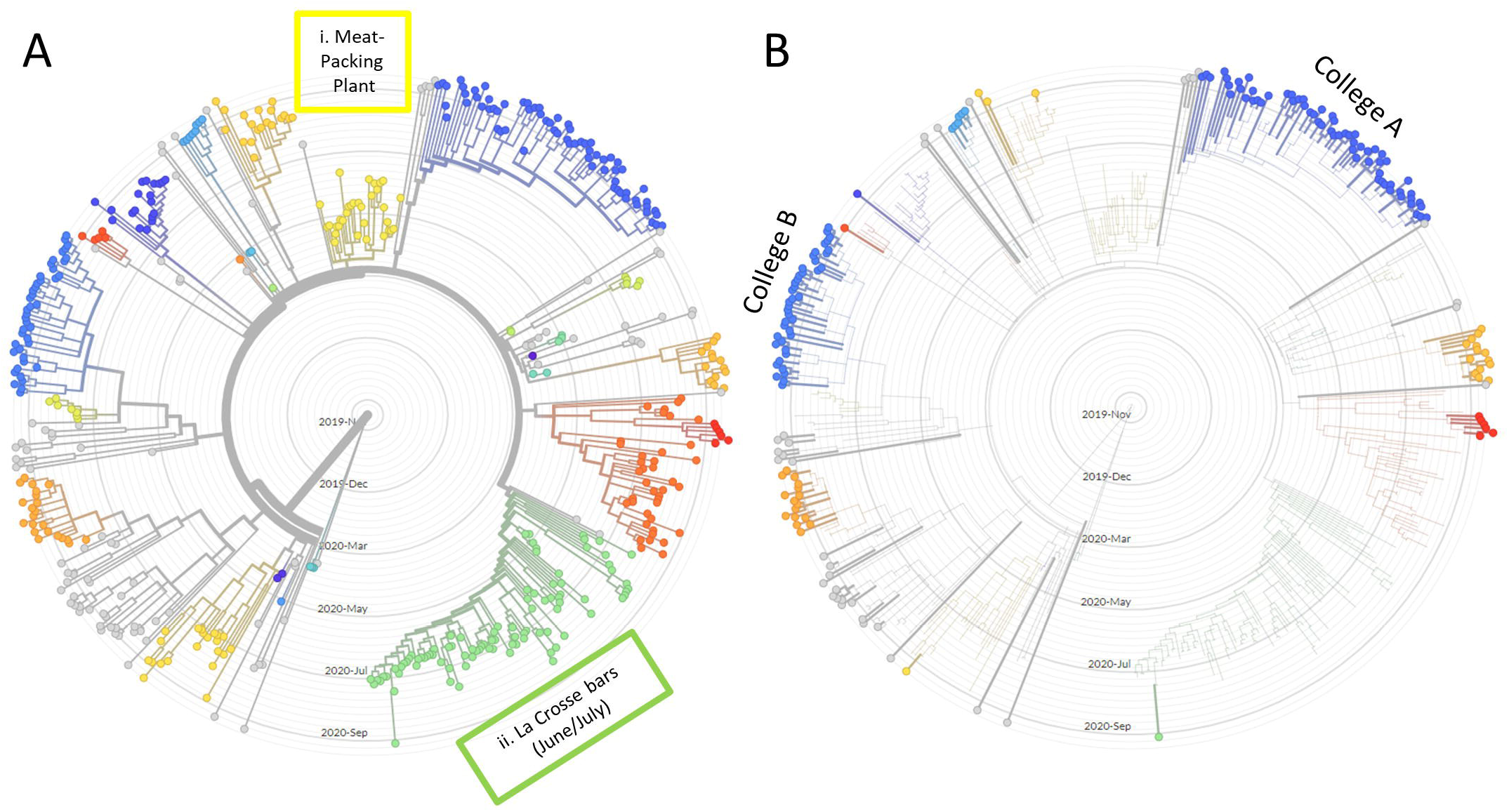
Radial time resolved phylogenetic trees containing (A) all 514 high-quality genomes sequenced between 3/18/20 and 9/28/20 and (B) highlighting just those specimens sequenced in September. Sequencing results are from a 21-county region of Wisconsin, Minnesota and Iowa. The case growth in La Crosse County in September was attributed to the College A and College B clusters.

During this period, all three academic institutions posted “COVID19 dashboards” on their websites, analysis of which indicated that the majority of cases were associated with the largest institution (University of Wisconsin – La Crosse, “UWL”), followed by Viterbo University and with relatively few cases affecting students enrolled at Western Technical College. UWL ran its own surveillance and diagnostic program for students living in dormitories (approximately 3000 students), but did not deploy resources sufficient to provide any surveillance for its large off-campus student population. Our sequencing program captured all positive tests conducted at the Gundersen Health System molecular diagnostic laboratory, a service availed of by many members of the off-campus population. Accordingly, while we generally lack genomes from the student dorm populations, our considerable coverage of the off-campus population and the presumably large amount of intermixing that occurred between the on and-off campus student populations due to the prevalent “party culture” (10,11) makes it unlikely that a substantial outbreak comprising an unrelated substrain might have occurred in and been restricted to the dorm-dwelling population.

Demographic details of the 111 individuals carrying the College A and B substrains are shown in Table 1. In total, 40.5% of the specimens were obtained from individuals aged between 17 and 24, with a further 16.2% from the 25-29 age group. For the entire cohort, 57.7% of cases affected females, although cases were more evenly distributed between sexes among the 17-29 year old group. 19 cases (17.1%) affected individuals aged over 60, including 4 cases among individuals in their 90s. This pattern of spread from clusters of younger age individuals into more vulnerable age groups is consistent with observations of a time-lag in which cases among older individuals tend to rise after outbreaks among young individuals (12). Although this statistical trend is evident from a nationwide analyses (12), our ability to establish direct genetic links using sequencing between the two age groups provides compelling evidence that risks of rapid spread of SARS-CoV-2 among college-age individuals are not limited to college environs but pose a direct threat to older persons in the surrounding community.

**Table 1:** Demographics of individuals associated with College A and College B clusters

|  |  | **College A (n=68)** | **College B (n=43)** | **Both Strains (n=111)** |
| --- | --- | --- | --- | --- |
| **Age Group** | **0-5** | 0 (0%) | 1 (2.3%) | 1 (0.9%) |
|  | **6-16** | 1 (1.5%) | 1 (2.3%) | 2 (1.8%) |
|  | **17-24** | 24 (35.3%) | 21 (48.8%) | 45 (40.5%) |
|  | **25-29** | 15 (22.1%) | 3 (7%) | 18 (16.2%) |
|  | **30-39** | 6 (8.8%) | 4 (9.3%) | 10 (9.0%) |
|  | **40-49** | 9 (13.2%) | 1 (2.3%) | 10 (9.0%) |
|  | **50-59** | 4 (5.9%) | 2 (4.7%) | 6 (5.4%) |
|  | **60-69** | 4 (5.9%) | 3 (7%) | 7 (6.3%) |
|  | **70-79** | 3 (4.4%) | 1 (2.3%) | 4 (3.6%) |
|  | **80-89** | 2 (4.7%) | 2 (4.7%) | 4 (3.6%) |
|  | **90-99** | 0 (0%) | 4 (9.3%) | 4 (3.6%) |
| **Sex (All ages)** | **Male** | 30 (44.1%) | 17 (39.5%) | 47 (42.3%) |
|  | **Female** | 38 (55/9%) | 26 (60.5%) | 64 (57.7%) |
| **Sex (Ages 17-29)** | **Male** | 22 (56.4%) | 10 (41.6%) | 32 (50.8%) |
|  | **Female** | 17 (43.6%) | 14 (58.3%) | 31 (49.2%) |

Detailed phylogenetic trees for both College A and B clusters are shown in Figure 4. Trees are scaled to show genetic divergence of the virus, such that samples aligned vertically are genetically identical and placement of a sample one increment to the right indicates acquisition of a single new mutation. Within both clusters, we found evidence of subgroups sharing identical viruses which typically reflect individuals infected in quick succession from a common source (e.g. at one or more parties). Groups of cases containing five or more genetically identical genomes are highlighted in orange, and the median age of individuals in these four groups were 20.5, 21, 25 and 29. This pattern is consistent with rapid spread of SARS-CoV-2 among the younger age group. Although both College A and B clusters included older individuals (See Table 1), analysis of the College B cluster indicated that this particular substrain had been transmitted into two skilled nursing facilities. The sequencing data revealed independent clusters affecting three patients in one facility and two staff and five patients in the second facility. Two of these patients died from COVID-19.

**Figure 4.**
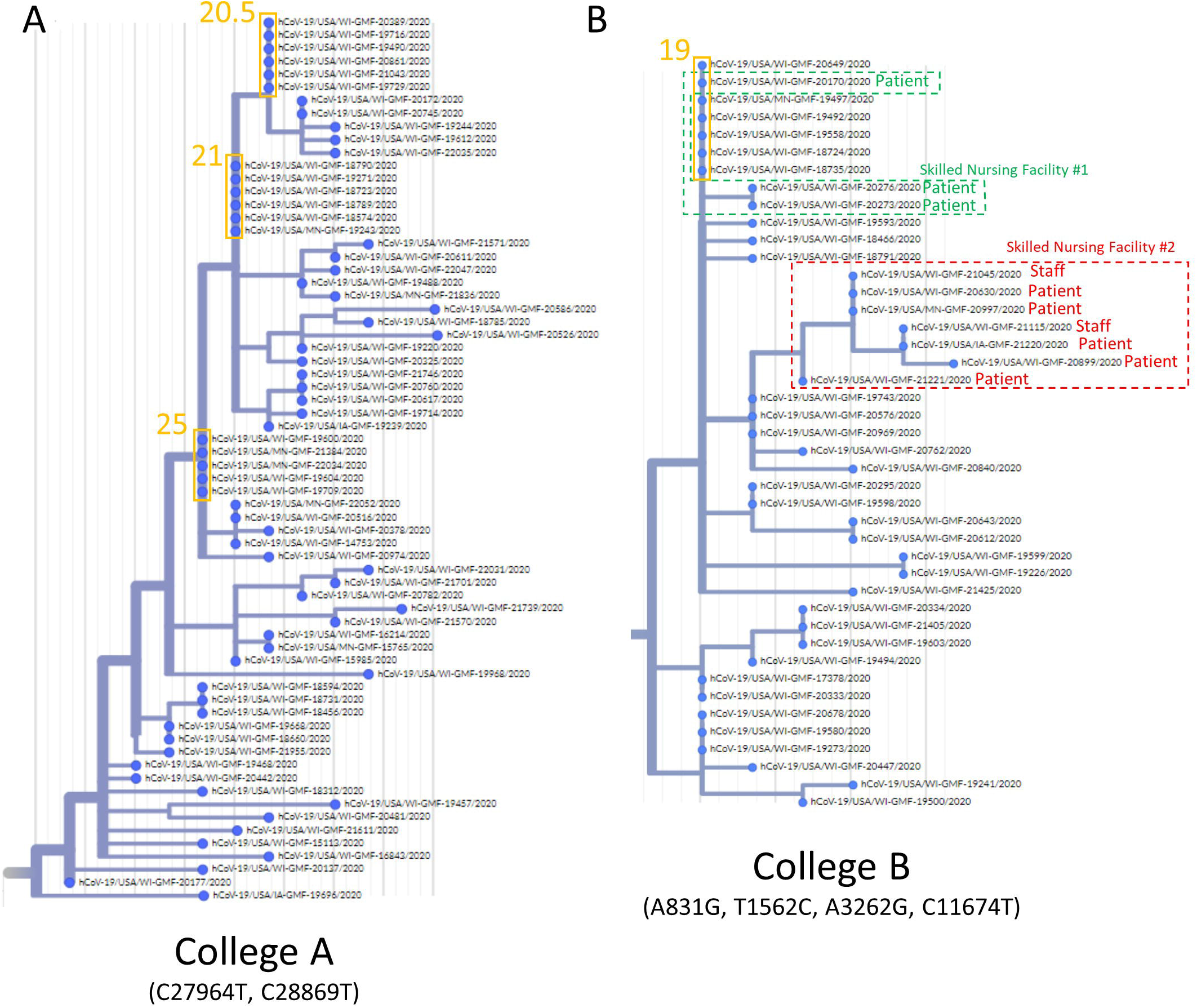
Phylogenetic trees demonstrating the relationships between cases in the College A and College B substrains. Trees are scaled by sequence divergence so that identical genomes are vertically aligned and each further increment to the right represents acquisition a new nucleotide variant. Clade-defining mutations are indicated in both cases. Groups of 5 or more genetically identical cases are highlighted in orange, with the median age of individuals in each group indicated. Transmission chains in two skilled nursing facilities, detected in the College B cluster, are also highlighted.

## DISCUSSION

This study provides an interim yet comprehensive look at a still-ongoing outbreak centered on the neighborhoods surrounding college campuses in a midwestern city, La Crosse, WI. While the 111 genomes sampled represent a fraction of the total cases in La Crosse county in this September outbreak, the data appear sufficient to draw some important conclusions about the nature of the outbreak and early consequences for vulnerable populations, as well as prompting consideration of whether various mitigation strategies adopted elsewhere could have been helpful.

Firstly, this large outbreak was overwhelmingly driven by just two substrains of SARS-CoV-2 which was quite surprising. Restarting of in-person education involved the return of many thousands of students from a wide range of cities and small towns throughout the Midwest at a time when SARS-CoV-2 cases in Midwestern 18-22 year olds were increasing at a rate more than twice the national average for that age group (13). Accordingly, we had expected greater viral diversity consistent with multiple independent viral introductions. In contrast, the major signal detected was for just two substrains which spread very rapidly among an at-risk population which socialized repeatedly at high-density alcohol-fueled indoor and outdoor gatherings. We cannot exclude the possibility that there were additional contemporaneous viral introductions and that these two substrains fortuitously happened to be the earliest to be exposed to large susceptible populations at events favoring rapid SARS-CoV-2 spread such that other potential substrains did not have the opportunity to access such a large susceptible population. Nevertheless, our data imply that the overwhelming majority of students were SARS-CoV-2-negative upon return to the campus environs and that rapid local spread, rather than imported cases, was the primary cause of the local outbreak.

Nationally, several universities made very considerable financial investments in pre-testing all students before their return to campus, a mitigation strategy that was not attempted in La Crosse. Our data suggest that, had that strategy been employed in La Crosse, it would have had a negligible impact on preventing the outbreak. At best, it might have delayed an inevitable outcome by a few days. Clearly, introduction of just two viral substrains was sufficient to fuel this very significant outbreak. In a setting where students from multiple colleges interact with each other and with other residents of a city with pre-existing SARS-COV-2 cases, student behavior will likely lead to significant outbreaks whether the initial student-imported viral input is large or small.

Testing of symptomatic and asymptomatic students after college opening is likely a more useful intervention, if performed at a sufficient frequency. When modeling multiple scenarios to mitigate SARS-CoV-2 spread during college re-opening for an 80-day fall semester, Paltiel and colleagues estimated that testing the entire student population every two days could limit cumulative infections to 4.9% of students, while weekly testing would result in 37% of students becoming infected (14). Importantly, both estimates involved assumptions of strong adherence to mask-wearing, handwashing, social-distancing, de-densifying classrooms and limiting bathroom sharing. The UWL plan for its 10,500 student body involved asymptomatic testing of its dormitory resident population (3000 students) once every 14 days with no surveillance testing among the much larger off-campus population. Considering that Paltiel estimated that testing 100% of an idealized guideline-adherent student population weekly would still lead to a cumulative infection rate of 37%, it seems likely that testing only 28% of an actual student population at half that frequency would be utterly inadequate to manage inevitable outbreaks. Success in Paltiel’s model also required efficiently functioning quarantine and isolation dormitories, a situation at variance with the chaotic situation at UWL where student reporters described individuals quarantined while the results of their tests were pending, partying in the quarantine dorm with COVID19-positive individuals, and then being released when their pre-quarantine test returned negative (15).

We detected these student-amplified infection clusters as part of a 21-county surveillance program in the Gundersen Healthcare System’s service area. Because this program was not student focused, we were collecting and sequencing other community cases in parallel. This allowed us to quickly detect the overspill from the college-age population in older adults. While these findings were consistent with public health expectations about risk to older population in settings of wide community spread and with epidemiological studies showing a statistical association in which case increases in young adults are typically followed by cases among older adults (12), our ability to genetically link these groups of cases provides direct evidence of transmission between these different age groups. Of particular concern, was the rapid transmission of one of these SARS-CoV-2 substrains into two skilled nursing facilities, causing sustained outbreaks with two fatalities so far. Our first case of what we came to call the “College B” cluster was collected on 8/27/20, a time when cases were slowly but steadily rising (Figure 1). The inflection point on the curve when cases began to rise more rapidly was on 9/10/20 (Figure 1) and the first skilled nursing facility patient specimen associated with this cluster was detected on 9/14/20.

## CONCLUSIONS

For college clusters, much concern has focused on higher risks to older faculty/staff as well as less frequent but significant adverse outcomes in younger individuals. In addition, university leaders clearly face a range of concerns including very significant financial pressures. Given the described testing strategy, the highly contagious properties of SARS-CoV-2, news reports of widespread outbreaks in other college communities that had attempted return to in-person instruction at earlier dates (16,17) and the well-understood risk-embracing nature of college students (18), the outbreak that occurred in La Crosse in September 2020 was likely inevitable once the decision to return to in-person instruction was taken. Our study highlights the very significant risks imposed by college administrator reopening decisions, not just on college-associated populations, but on vulnerable individuals in surrounding communities. Although our primary focus has been on using viral genetics to track introduction and spread of these substrains, we note that the adverse effects of college-amplified clusters are not limited to the morbidity and mortality of infected individuals and the substantial associated healthcare costs. Rapid case growth, as experienced in La Crosse, intensifies the magnitude of the multifaceted adverse effects of the pandemic upon communities. These effects include significantly extending the period during which children are unable to attend in-person K-12 schooling (19), further exacerbation of the many psychosocial stresses affecting both individuals and families (20), extending the ongoing unemployment crisis (21), placing more families at risk of losing health insurance (22) and harming small businesses (23). These factors should all be considered as part of any cost-benefit analysis of reopening colleges and universities.

## Data Availability

All genomic data have been deposited in GISAID.

https://www.gisaid.org/

## ACKNOWLEDGEMENTS

This work was supported by the Gundersen Medical Foundation. PK holds the Dr. Jon & Betty Kabara Endowed Chair in Precision Oncology. We thank the members of the Gundersen Medical Foundation’s Molecular Diagnostics Laboratory for providing the specimens used in this study.

